# Spatio-temporal modelling of COVID-19 infection and associated risk factors in Dakar, Senegal

**DOI:** 10.1101/2025.07.04.25330897

**Authors:** Assane N. Gadiaga, Mame Wodji Tine, Aminata Niang Diene, Catherine Linard, Niko Speybroeck, Ortis Yankey, Somnath Chaudhuri, Chibuzor Christopher Nnanatu, Eimear Cleary, Shengjie Lai, Attila N. Lazar, Andrew J. Tatem

## Abstract

The spread of infectious diseases is a major threat to global health and economy and the recent COVID-19 pandemic is a perfect illustration of this. Appropriately modelling and accurate prediction of the outcome of disease spread over time and across space is a critical step towards informed development of effective strategies for public health interventions. In low and middle-income countries, however, the scarcity of spatially disaggregated time-series infectious diseases data often limits the analysis of the burden of infectious disease at a broad-scale, and the effects of the contextual risk factors is not often fully captured. In this study, we investigate the spatiotemporal patterns of COVID-19 infection in Dakar at the neighbourhood level, and evaluate the impact of potential risk factors. Geostatistical models based on COVID-19 infection were used to explain and predict the spatiotemporal distribution of COVID-19 infection between June 2020 and June 2021. We specified a Bayesian regression model that incorporates a spatio-temporally autocorrelated random effect in order to quantify the evolution of the spatial patterns of the COVID-19 infection overtime. Results show significant strong spatial heterogeneity but relatively small temporal variations of the COVID 19 distribution, and a positive association between adjusted population density (mean of the posterior probability: 0.29, credible interval: 0.24-0.34) and residential areas (mean of the posterior probability: 1.25, credible interval: 0.66-1.83) with COVID-19 infection. Western areas are at higher risk of COVID-19 infection compared to eastern and less densely populated peripheral neighbourhoods. Measuring the role of contextual risk factors and mapping the at-risk areas can provide valuable insights for policymakers in low- and middle-income countries, enabling more targeted public health interventions. These efforts also support the management of endemic diseases and preparedness for future outbreaks.

## 1. Introduction

The COVID-19 pandemic has had profound health, economic, and social consequences worldwide throughout the year 2020 until now. Since its emergence in late 2019, the disease has spread rapidly, leading to unprecedented public health responses such as lockdowns, social distancing measures, and mass vaccination campaigns [1]. As of April 2024, the COVID-19 related death toll had risen to more than 7 million from more than 700 million reported infections [2]. While widespread vaccination efforts have significantly reduced transmission and mortality [3], the containment of the virus is challenging because of the biological features of the virus [4], involving mutating variants that mitigate the effectiveness of the vaccine, leading to sustained rates of infection throughout the community.

Given the persistent risk of COVID-19 variants worldwide, assessing the disease spread and understanding its spatiotemporal patterns are key to effective monitoring, prevention and control. This is particularly true in many LMICs, where fragile disease surveillance system were unable to monitor the progress of COVID-19 and formulate appropriate responses. Recent studies have explored the role of various contextual factors in shaping the spread of COVID-19, including population density, landscape connectivity, human mobility, socioeconomic characteristics, environmental conditions, pollution and smoking [5–11]. Population density, in particular, has been widely investigated as a risk factor of disease transmission, given the fact that people’s proximity to each other increases likelihood of infection [12]. In many cases, the relationship was positive, with more densely populated areas having higher rates of infections or experiencing early outbreaks [5,10,13,14]. Highly connected areas also face earlier and higher COVID 19 infection rates [15–18], and socioeconomic factors such as residential characteristics drive disparities in COVID 19 cases and deaths [7,19–21].

Several statistical approaches have been used to investigate the role of the aforementioned risk factors on the spread of the COVID-19 disease. They are either classical and frequentist methods, which remain at the core of most practical statistical applications [22], or Bayesian methods, which have been increasingly used in the field of spatial epidemiology due to their ability to incorporate spatial and temporal dependencies [23,24].

Although the scientific community is deploying efforts to understand contextual factors invloved in the spread of COVID-19, existing epidemiological data are not often resolved at spatial levels that allows for in-depth analyses; they are in most cases available at a large scale (i.e., regional or national levels) and the patterns of the disease at finer levels (i.e., at the intra urban levels) remains unknown.

In Senegal, the first case of COVID-19 was reported in early March 2020, and the majority of subsequent Senegalese cases were reported in Dakar, the capital city. According to the Ministry of Health, as of May 29, 2020, the Dakar region accounted for nearly three quarter (2525 cases out of the 3429 detected in the country; 73.6%) of reported COVID-19 infections in Senegal [25]. Public health authorities classified the source of the COVID-19 infection as one of the following four categories: (1) contact cases; (2) community cases; (3) imported cases and (4) evacuated cases. An infection was termed as a community case when the source of infection was unknown [26], making efforts to control the pandemic more challenging. Against this backdrop, the present study analysed the spatiotemporal dynamics of COVID-19 disease in Dakar, with a focus on the COVID-19 infection which arose from community transmission, and using COVID-19 time-series data at the finest administrative level.

Through a spatio-temporal Bayesian framework we evaluate the impact of population density, temperature, distance to marketplaces, and residential characteristics on COVID-19 spread. Distance to marketplaces is used here as a proxy of mobility [27], while residential and non-residential areas indicates differences in living conditions and is a useful proxy on poverty levels [28]. In addition to the widely used population density (count of people per total area), adjusted population density that refers to the count of people per built-up area was also considered. This approach seeks to better capture the effect of crowding, because, although density expresses a ratio between population and an unit of area, these units are not uniformly distributed over the space. Our aim is to understand the spatiotemporal dynamic of COVID-19 spread in Dakar as well as related potential drivers. In a context of fragile health system, as in many low- and middle-income countries, having detailed time-series epidemiological data at the finest administrative level for Dakar offers a unique opportunity to explore advanced spatial statistical modelling approach and improve our understanding of localised transmission patterns. Understanding these localised transmission patterns is essential for designing targeted public health interventions, optimizing resource allocation, and mitigating the impact of future outbreaks.

## 2. Materials and methods

### 2.1. Study area and data

#### 2.1.1. Study area

Dakar is the capital city of Senegal (Fig 1) and an example of African city that combines several aspects of the Africa’s urban growth: rapid urbanization, land cover and land use changes, inequalities in living conditions [29] that have led to different health status [28]. Located in the westernmost part of the African continent (Fig 1), Dakar is the driving force of the Senegalese economy and through its markets, banks and industrial sector remains a magnet for migrants from all over the country. According to the last national census conducted in 2023, the region had a population of 4 million inhabitants, nearly a quarter of the Senegalese population (22 %) in an area representing only 0.3 % of the country [30]. The availability of georeferenced fine scale census information [28] and high quality, very-high resolution remote sensing land cover and land use products [31,32] has improved our knowledge on health inequalities and associated socioeconomic and environmental risk factors [28,33].

**Fig 1.**
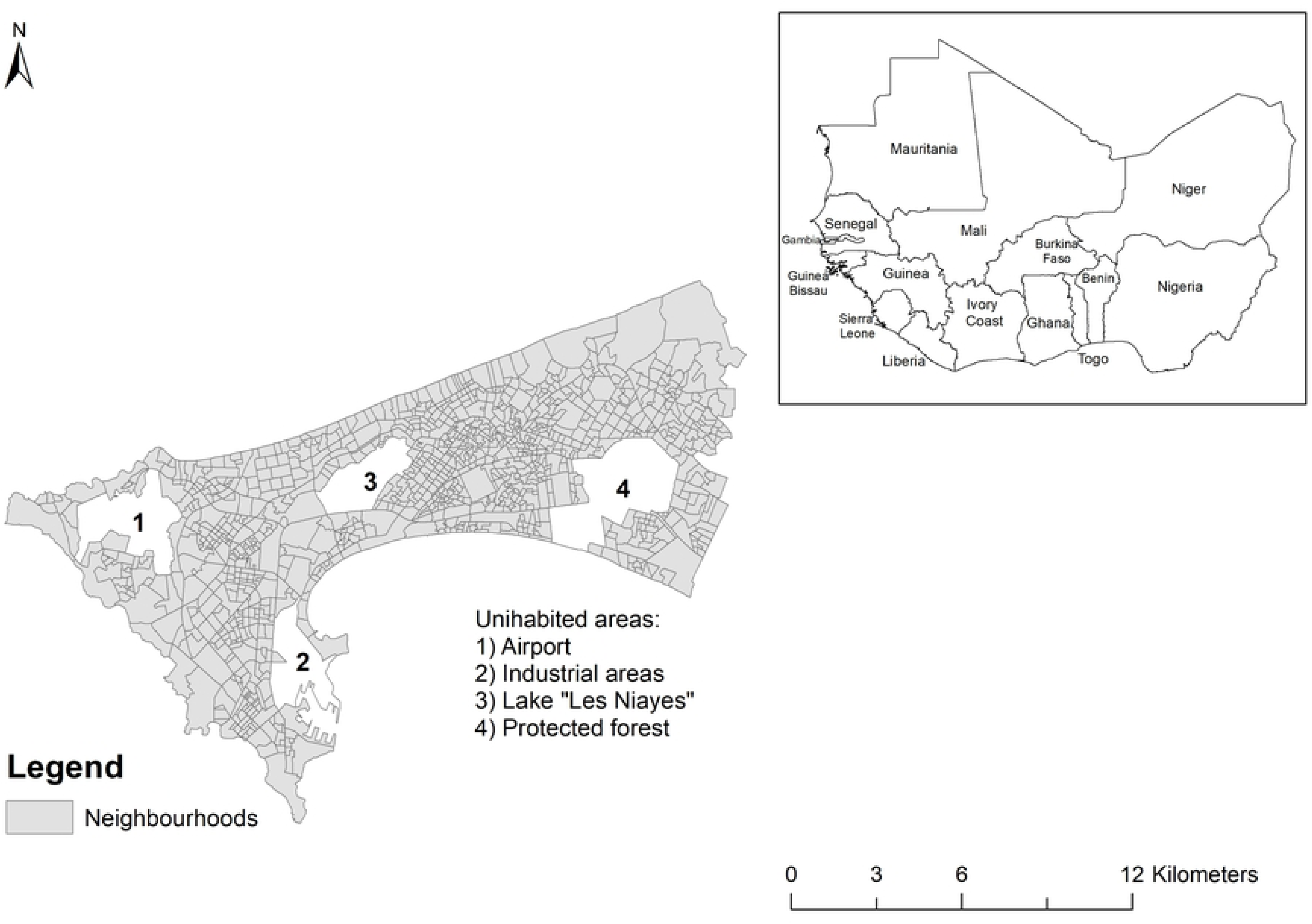
Geographical location of Dakar.

#### 2.1.2. Data

The epidemiological data used in this study were obtained from the daily reports of COVID-19 infection in Dakar produced by the Ministry of health [25]. Data cover the period from 01/06/2020 to 30/06/2021 and records the number of confirmed COVID-19 infections from community-based transmission. Each anonymised record includes the date of the confirmed infection, the location of the infection and the number of new infections. There can be more than one record per date, with similar or different locations. The location of the infection often refers to the neighbourhood, which is the smallest administrative unit in an urban area. However, in some situations, the location can refer to a higher administrative extent, i.e. the commune, or a lower level spatial extent such as the housing unit. Geographic coordinates of each infection record were obtained by joining the excel-based disease data file with the neighbourhood shapefile. To do this, the unique identifier availabe in the latter was assigned to the disease record when there is a matching between the location of the infection and the polygon shapefile. When the location of the infection was referring to a Commune or a housing unit, a fuzzy-matching algorithm was used to merge the infection record with the neighbourhood shapefile. After the geocoding of the infection records, the daily cases were aggregated by month for each neighbourhood. Thus, each neighbourhood has an aggregated count spanning the study period, providing an idea of the basic severity status of the neighborhood during the pandemic <u>(covid19_Dakar)</u>. Although the neighborhood-level data offer an opportunity to examine the intra-urban variation of COVID-19 infection in Dakar, it is important to note that this does not provide a complete picture of the status of the pandemic in the city, as only infections from community transmission are analysed. In fact, only community based transmission can be used to reconstruct the spatio-temporal dynamic of the pandemic because, unlike the other type of infection, it includes information about the location of the infection (patient’s residence).

Furthermore, just as there are multiple records for some dates, there are also days with no reported infections from community transmission. This might weaken the quality of the study, although it remains unclear whether the absence of reported infections is due to a lack of such cases among tested patients on the corresponding dates. The cumulative incidence and temporal distribution of COVID-19 infections over the study period are illustrated in Figs 2 and 3, respectively.

**Fig 2.**
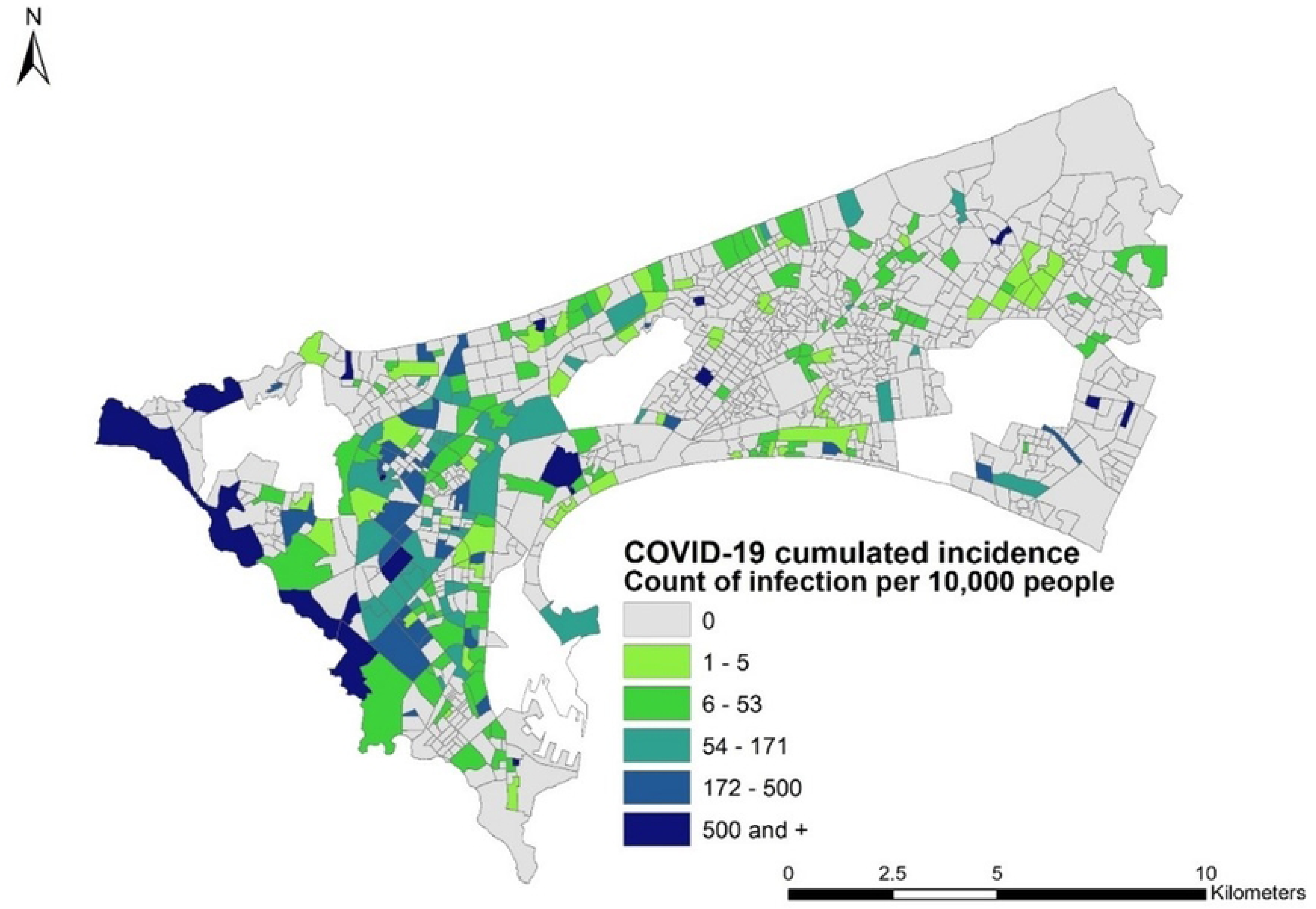
Aggregated incidence of COVID-19 confirmed infection from community transmission from June 2020 to June 2021.

**Fig 3.**
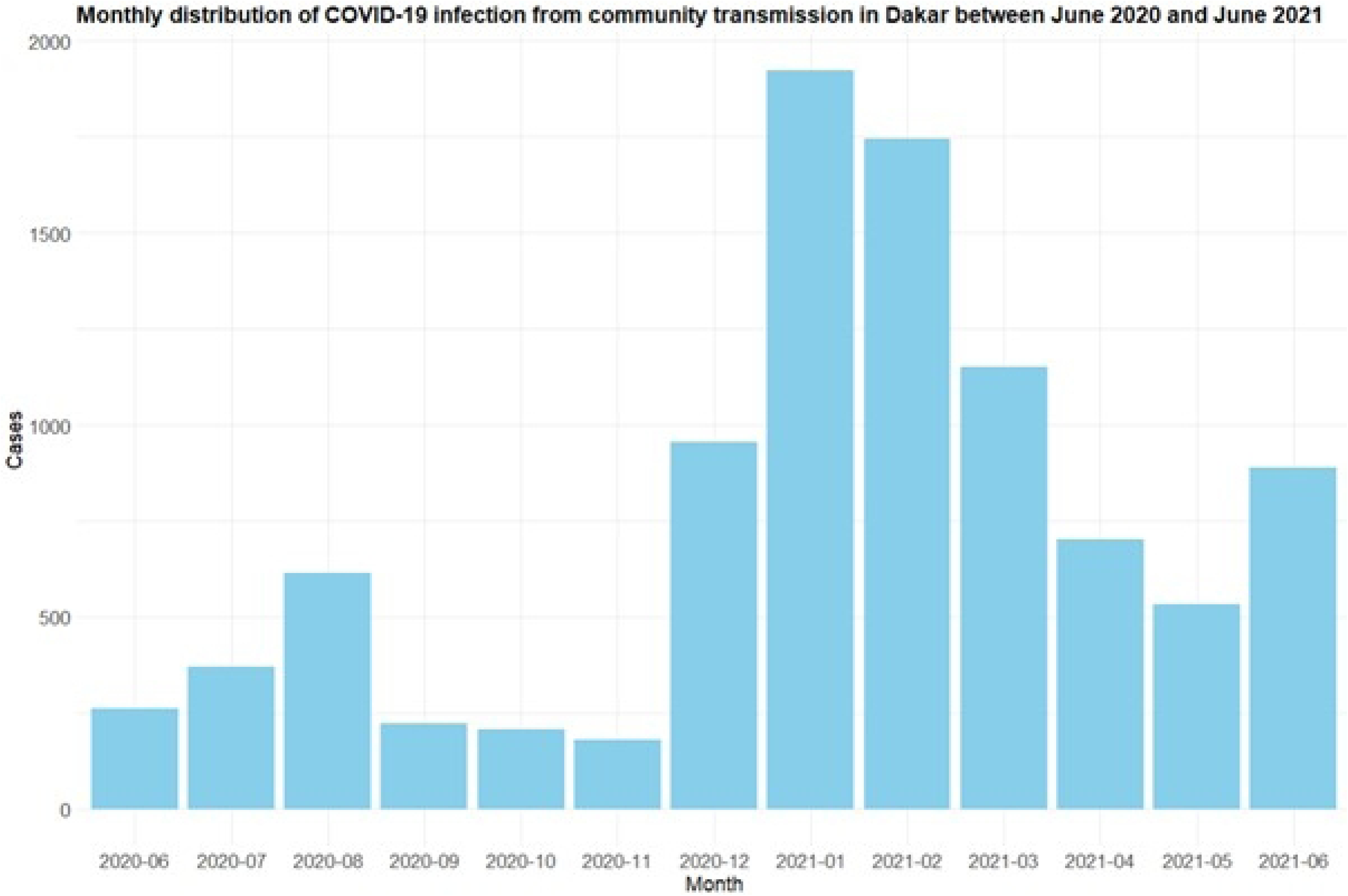
Monthly distribution of COVID-19 infection from community transmission in Dakar from June 2020 to June 2021.

As predictors of COVID-19 infection, we used two high resolution satellite derived land use [32] and gridded population datasets [34], with a spatial resolution of 10m for the population data and street block level for the land use map. Additionally, distance to marketplaces and temperature data obtained from the WorldPop data repository were used [35]. The land use map depicts several spatial metrics, such as residential built-up areas (planned or unplanned), non-residential built-up areas (administrative, commercial and service areas), agricultural vegetation, natural vegetation, water bodies and baresoil areas. It is a precise representation of the Dakar’s urban landscape, with an overall accuracy of 79 % [32] and regarding its usefulness in understanding inequalities in health and living conditions [28,33]. In the present study, however, this accuracy may be hampered by the mismatch between the year the satellite data were collected (2015) and the year in which the COVID-19 data were obtained (2020-2021). Although built and settlement layers are time-variant, we assume that this 5 year gap is negligible in the particular context of Dakar. To further assess this, we used the world settlement footprint data [36] and calculated the average percentage of change in the built-up areas at the neighbourhood level from 2015 to 2019. On average, built-up areas per neighbourhood have grown by 5% within the period 2015-2019 in Dakar, indicating minimal changes. It is important to note that the land use data does not cover Goree and Ngor islands as well as some parts of eastern Dakar. Thus, to ensure a full overlap between the COVID-19 data and the geospatial predictors, the spatial extent of the study area was reduced and sparsely built-up areas in the eastern urbanisation front were excluded from the analysis.

Using these remotely sensed data sources, we first extracted the average residential built-up areas, non-residential built-up areas, temperature, and distance to marketplaces for each neighborhood. The total gridded population per neighborhood was divided by the neighborhood’s total area to calculate population density. We then computed the adjusted population density by dividing the gridded population counts by the total built-up area for each neighborhood. Additionally, built-up area was divided by the neighborhood’s total area to calculate built-up density. The covariates used for the modeling are listed in Table 1.

**Table 1.** List of covariates used for the modelling.

| Variable | Description | Source |
| --- | --- | --- |
| Temperature | Annual average temperature | WorldPop |
| Built-up density | Built-up area per total area | MAUPP<sup>a</sup> |
| Distance to marketplaces | Average distance to marketplaces | WorldPop |
| Population density | Count of people per total area | MAUPP |
| Adjusted population density | Count of people per built-up area | MAUPP |
| Residential areas | Proportion of land use (LU) used for residential purposes | MAUPP |
| Non-residential ACS areas | Proportion of LU without residences used for administrative, commercial and services (ACS) purposes | MAUPP |

<sup>a</sup>MAUPP = Modelling and forecasting African Urban Population Patterns for vulnerability and health assessments (https://maupp.ulb.ac.be/).

Statistical analyses were performed to check for multicollinearity among the covariates and to remove any covariates that were not statistically significant. To address this, we used the Variance Inflation Factor (VIF) to measure the level of collinearity. Covariates with a VIF greater than 5 were considered to have high multicollinearity and were removed from the analysis [37,38]. Only covariates with a VIF less than 5 and those that were statistically significant were selected for the final model. Among the 7 variables considered for this analysis and introduced in Table 1, only built-up density was found showing multicollinearity issue and further dropped. In the following section, the Bayesian statistical modelling approach we developed to conduct this research is presented.

#### 2.1.3. Ethics statements

The epidemiological datasets used in this study were completely anonymised and aggregated to administrative level to avoid confidentiality issues in accordance with relevant data protection regulations. The study adhered to the Declaration of Helsinki as it ensures respect for all participants and protect their privacy, health and rights. Additionally, the data has been made publicly available in the official website of the Ministry of Health through a web-based visulisation platform.

### 2.2. Modelling COVID-19 infection risk

The spread of infectious diseases like COVID-19, involves complex patterns over both space and time. This complexity arises from the interactions between human behaviour, environmental factors, and disease transmission. To analyse COVID-19 cases in Dakar, Senegal, we have used a Bayesian spatiotemporal modelling approach. This method allows us to account for both spatial and temporal autocorrelation structures in the data. Spatial autocorrelation refers to the way disease cases are clustered in certain areas, while temporal autocorrelation deals with how cases change over time. Our modelling approach is important because COVID-19 cases are not evenly distributed across different locations or time periods, as illustrated in Figs 2 and 3.

In this study, we applied three distinct modelling approaches to analyse the spread of COVID-19 in Dakar. Our first approach focused on identifying monthly trends across the entire city. For this, we used both first-order and second-order random walk models (RW1 and RW2) to capture temporal randomness. To account for neighbourhood-level variations, we included an independent and identically distributed (IID) random effect. In this approach, we aggregated the total number of COVID-19 cases across Dakar for each month, using these monthly totals as the response variable.

In the second approach, we examined COVID-19 case counts at the neighbourhood level, capturing spatial variations within Dakar in addition to temporal trends. This separable spatiotemporal model included spatial randomness based on individual neighbourhoods, temporal randomness, and the IID effect of each neighbourhood. Finally, our third approach also involved spatiotemporal modelling, but with an added spatiotemporal interaction effect. This term represented an unobserved independent effect for each combination of neighbourhood and month, allowing us to capture unique patterns in each area over time.

#### 2.2.1. Temporal modelling of COVID-19 infection

In the first approach, we have employed a temporal Poisson regression framework to analyse the temporal dynamics of COVID-19 in Dakar. In this model, the number of confirmed cases at each time point was represented using a Poisson likelihood in that we are dealing with a count data. The temporal resolution of the analysis has been set to one month, allowing us to explore trends and changes in the number of infected cases over time. Additionally, we incorporated geospatial variables to assess their potential correlations with the infection rates.

Let *Y*_*t*_ and *E*_*t*_ represent the number of observed and expected COVID-19 cases in the population at time *t*, where *t* = 1,…,*T*. We assume that, given the relative risk *q*_*t*_ the number of observed cases follows a Poisson distribution:

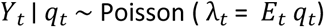

where *E*_*t*_ is the total population for the study area, and the log-risk is modelled as:

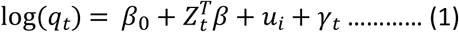

where, *Z*_*t*_ represents the matrix of covariates used to account for various factors influencing the infection rates. The term *u*_*i*_ is an IID random effect that captures unobserved variability across the neighbourhoods. Additionally, γ_*t*_ represents the temporal effects, which can be modelled using either a RW1 or a RW2 to account for the temporal structure and potential trends over time. In the case of the RW1, the temporal effects γ_*t*_ are modeled such that the increments Δγ_*t*_ = γ_*t*_ ― γ_*t*―1_ follow a Gaussian distribution with zero mean and precision τ. On the other hand, for a RW2, the temporal effects are modeled as Δ^2^ γ_*t*_ = γ_*t*_ ―2 γ_*t*―1_ + γ_*t*―2_ also follow a Gaussian distribution with zero mean and precision τ. This RW2 model provides a smoother temporal evolution over time compared to the RW1 model, making it particularly useful for capturing complex temporal patterns such as trends, cycles, or deviations from simple linear temporal patterns. For prior selection, we have used penalized complexity (PC) priors [39]. For the IID random effect, the PC prior on the precision was set to [0.5, 0.01], where 0.5 is the central tendency and 0.01 controls the spread or uncertainty around it. For the RW1 or RW2 model on the monthly data, the PC prior on the precision with parameters [1, 0.01] represents a location parameter of 1 and a scale parameter of 0.01, indicating a prior belief that the precision is centered around 1 with relatively low uncertainty. The prior for the coefficient vector β = (β_0_,…,β_*p*_) which follows a zero-mean Gaussian distribution with a precision of 0.001. All log-precision parameters followed inverse gamma distributions with shape parameter (α = 1) and scale parameter (β = 0.00005).

#### 2.2.2. Spatiotemporal modelling of COVID-19 infection

In the second approach, we employed a spatiotemporal Poisson regression framework to analyze the spread of COVID-19 in Dakar. In this model, the number of confirmed cases at each spatial unit (neighborhood) and time point was represented using a Poisson likelihood. Similar to the first approach, the temporal resolution of the spatiotemporal analysis was set to one month, allowing us to observe trends and changes in case numbers over time (months). The spatiotemporal model included spatial random effects to account for the influence of neighboring regions on case numbers. It also incorporated temporal random effects to capture trends and seasonal variations over time. Additionally, geospatial covariates were included to explore their potential associations with infection rates.

Let *Y*_*it*_ and *E*_*it*_ represent the number of observed and expected COVID-19 infected cases in the *i*-th area (neighbourhood) and the *t*-th period (month), where *t* = 1,…,*T*. We assume that, given the relative risk *q*_*it*_, the number of observed cases follows a Poisson distribution:

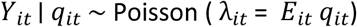

where, *E*_*it*_ are the total population for individual neighbourhood of the study area and the log-risk is modelled as in Equation (2) for spatiotemporal separable model and in Equation (3) for spatiotemporal interaction model.

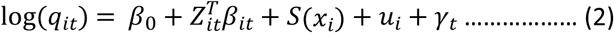

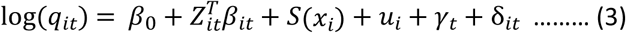

where in both Equation (2) and (3), *S*(.) represents the spatially structured random effects, and *Z*_*it*_ represents the matrix of covariates used in the model. The temporally structured effect has been represented by γ_*t*_. In Equation (3), δ_*it*_ represents the spatio-temporal interaction between the two structured effects. The IID random effect *u*_*i*_ is added to the model to capture neighbourhood-level variability with a penalized complexity (PC) prior on the precision set as [0.5,0.01] where we assumed a prior mean of 0.5 and a prior precision of 0.01. We have modelled temporal trends using a RW1 for the monthly data. This model was scaled for identifiability, and we applied a PC prior on the precision with parameters [1,0.01] with a prior mean of 1 and a prior precision of 0.01. For all prior selections, we used PC priors following the framework proposed by Simpson et al. [39]. We have assigned a vague prior to the vector of coefficients β = (β_0_,…,β_*p*_) which follows a zero-mean Gaussian distribution with a precision of 0.001. Finally, all parameters associated with log-precisions are assigned inverse Gamma distributions with shape parameter (α = 1) and scale parameter (β = 0.00005). The default prior distributions for all parameters in R-INLA were selected based on commonly used priors in previous studies [40–42]. Our findings demonstrate robustness to alternative priors. We have tested multiple scenarios with different priors and consistently obtained the same results. As shown in Equation (3), we introduced a spatiotemporal interaction effect as an unobserved, independent term for each region-period combination (*i*,*t*), modelled as 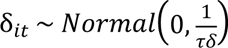,. This interaction term assumes no additional structure, representing independent variations from the main spatial and temporal effects when those are included in the model. It captures global space-time heterogeneity and is typically modeled as white noise, as discussed in previous studies [40]. This interaction term offers a more flexible structure, capturing localized deviations from the main spatial and temporal patterns. To compare modelling strategies, we constructed the third model set that incorporate all covariates, using both spatiotemporal interactions and PC priors. A summary of these competing models is presented in Table 2 in the Results Section.

**Table 2.** Posterior estimates of model parameters of temporal and spatiotemporal models of COVID-19 in Dakar.

|  |  |  | Spatiotemporal Model |  |  |  |
| --- | --- | --- | --- | --- | --- | --- |
|  | Temporal Model (M1) |  | Separable Model (M2) |  | Interaction model (M3) |  |
| Variable | Mean | 95% BCI <sup>a</sup> | Mean | 95% BCI | Mean | 95% BCI |
| Intercept | -10.23 | -55.38,-34.91 | -10.25 | -55.42,34.92 | -10.27 | -55.46,34.92 |
| Residential areas | 1.25 | 0.67,1.83 | 1.25 | 0.67,1.83 | 1.25 | 0.66,1.83 |
| Non-residential areas | -0.66 | -1.29,-0.04 | -0.66 | -1.29,-0.04 | -0.67 | -1.29,-0.04 |
| Population density | -0.003 | -0.003,-0.002 | -0.003 | -0.003,-0.002 | -0.003 | -0.003,-0.002 |
| Adjusted population density | 0.29 | 0.24,0.34 | 0.29 | 0.24,0.34 | 0.29 | 0.24,0.34 |
| Temperature | 0.03 | -0.12,-0.18 | 0.03 | -0.12,0.18 | 0.03 | -0.12,0.18 |
| **Distance to marketplaces** | 0.00 | 0.00,0.00 | 0.00 | 0.00,0.00 | 0.00 | 0.00,0.00 |
| **DIC<sup>b</sup>** | 37044 |  | 37043 |  | 36711 |  |
| **WAIC<sup>c</sup>** | 37056 |  | 37055 |  | 36885 |  |

<sup>a</sup> BCI: Bayesian Credible Interval
<sup>b</sup> DIC: Deviance Information Criterion
<sup>c</sup> WAIC: Watanabe-Akaike Information Criterion

The dataset used in this study shows zero inflation. As such, zero-inflated Poisson and Poisson Hurdle models can be used to accommodate zero-inflated discrete distributions. While ZIP models add extra probability mass to the zero outcome [43], Poisson hurdle models, on the other hand, are characterized by a mixture of zero and non-zero outcomes [44,45]. In a ZIP model, the response variable *Y*_*it*_ is zero with probability π, and follows a Poisson distribution with parameter λ_*it*_ with probability 1 ― π. In particular,

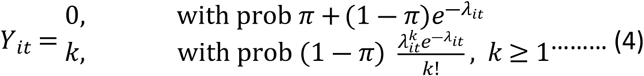

Conversely, in a Poisson hurdle model, the response variable *Y*_*it*_ is zero with probability π, and follows a truncated Poisson distribution with parameter λ_*it*_ with probability 1 ― π. Mathematically,

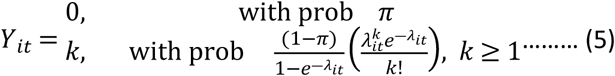

In the model fitting process, Bayesian inference typically employs Markov Chain Monte Carlo (MCMC) methods to estimate the joint posterior distribution of model parameters [46,47]. But for big dataset such as the time-series data used in this study, this can be computationally intensive. As a faster alternative, the integrated nested Laplace approximation (INLA) approximates posterior marginal distributions efficiently, especially for models represented as latent Gaussian Markov random fields (GMRF) [42,48]. In the current study, we have applied INLA along with a stochastic partial differential equation (SPDE) method. The SPDE method allows us to model a continuous spatial process, such as a Gaussian field (GF), using a discretely indexed spatial random process like a GMRF [49,50].

In particular, the spatial random process represented by *S*(.) explicitly denote dependence on the spatial field, follows a zero-mean Gaussian process with Matérn covariance function [51] represented as:

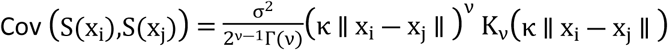

The parameter σ^2^represents the process variance, indicating the variability of the values around the mean. The smoothness parameter *v* > 0 influences the rate of decay of correlations with distance, while the scaling parameter *κ* > 0 governs the overall range of the covariance. The term ∥(x_i_ ― x_j_ ∥) denotes the Euclidean distance between locations. The modified Bessel function of the second kind, *K*_*v*_(.), defines the covariance structure, and Γ(*v*) is the gamma function, normalizing the covariance for scaling purposes.

The parameterized model we follow is of the form:

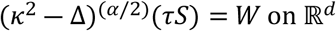

where 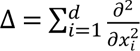 is the Laplacian, α = (*v* + *d*/2) is the smoothness parameter, τ is inversely proportional to σ, *W* is Gaussian white noise and *κ* > 0 is the scale parameter, related to range *r*, defined as the distance at which the spatial correlation becomes negligible. For each *v*, we have *r* = 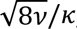, with *r* corresponding to the distance where the spatial correlation is close to 0.1. Note that we have *d* = 2 for a two-dimensional process, and we fix *v* = 1, so that α = 2 in this case [40–42]. In the current study, we have applied the PC priors [39] for the Matérn model, which helps prevent overfitting by supporting simpler models. The prior for the marginal standard deviation is set as [1,0.1] where we assumed a prior mean of 1 and a prior precision of 0.1. This allows some flexibility around a mean of 1 based on our expectations of spatial variation. We utilized the centroids of each neighborhood as the target locations for SPDE mesh construction. According to Verdoy [52], an optimal mesh should possess a sufficient number of vertices to facilitate effective predictions, and within a limit to have control over computational time. Following this principle, we selected the best-fitting mesh having 7365 vertices. S1 Fig depicts the SPDE mesh designed for the study area with 1078 neighborhood centroid locations highlighted as red points and the blue line marks the boundary of the study area. We have created a projection matrix to transfer the spatially continuous Gaussian random field from the observations to the mesh nodes. The centroids of the neighbourhoods and the triangulation in the mesh are used to develop this projection matrix.

In the model fitting process, we have included seven covariates, such as population density, adjusted population density, built-up density, average value of temperature, residential built-up areas, non-residential built-up areas, distance to marketplaces, as outlined in Section 2.1. The variable month spans from 1 to 12, representing each month of the year, and month is treated as factor in the model. We used data on COVID-19 cases from June 2020 to May 2021 (a total of 12 months) to fit the model described in Equation (2). To assess prediction accuracy, we held out data from the following month, June 2021, as a test set. Using this fitted model, INLA then predicted the COVID-19 cases for these held-out months by computing the predictive distributions based on the posterior distributions of the model parameters and latent variables. This approach allows INLA to provide both point predictions and uncertainty estimates, making it suitable for evaluating model accuracy on out of sample data. Finally, we compared the predicted values with the original observed COVID-19 cases from June 2021 to evaluate the predictive performance of the model, with detailed comparison results and performance metrics discussed in Section 3. All the analyses were conducted in R statistical software (v.4.2.2).

## 3. Results

The following results summarize the outcomes of our Bayesian spatiotemporal modelling approaches, highlighting the temporal dynamics, spatial heterogeneity, and zero-inflation characteristics of COVID-19 case distributions in Dakar. For the latter, a ZIP hurdle distribution is used in the modelling, and the parameter significance as well as the goodness of fit of the three fitted Bayesian ZIP hurdle models are presented in Table 2. It appears that the spatiotemporal interaction model was the best model to fit the data. Indeed, the best fitting spatiotemporal interaction model (M3) has the lowest Deviance Information Criterion (DIC) and Watanabe-Akaike information Criterion (WAIC) values compared to the temporal (M1) and separable spatiotemporal model (M2). The DIC and WAIC values varied slightly from the temporal to the separable spatial-temporal models (37050 and 37049 DIC values for M1 and M2, respectively, then 37061 and 37060 WAIC values for M1 and M2, respectively). When a spatiotemporal interaction is specified, the DIC and WAIC has significantly dropped, indicating higher predictability when the space-time interaction is accounted for.

For the models specified in Equations (2) and (3), we examine the posterior distributions of fixed and random effects for both the separable spatiotemporal model and the spatiotemporal interaction model. These posterior distributions are presented in S2 and S3 Figs, and S4 and S5 Figs. For both models, the posterior distribution of the fixed effects provides insights into the impact of covariates on the outcome across space and time.

The results indicate that the covariates month number, distance to marketplace, and temperature are non-significant in both models, with null posterior mean or zero included within the range of the 95% credible interval. Conversely, population density and non-residential ACS areas show a significant negative effect, suggesting a negative association in the model. Adjusted population density and residential areas exhibit a significant positive effect, indicating a strong positive influence. For both spatiotemporal models, the fixed effects demonstrate a similar trend and nature.

On the other hand, the posterior distribution of the random effects captures underlying spatial, temporal, and spatiotemporal variability. In the separable model, spatial and temporal effects are modeled independently, assuming no interaction between them. In contrast, the spatiotemporal interaction model includes an additional interaction term, allowing for unique variations in each region-period combination. S2 Fig depicts the marginal posterior mean of spatial, IID and temporal random effects with 95% credible intervals. The random effects analysis indicates that the IID random effect (neigh_id_index) has a stronger influence compared to the spatial random effect (denoted as i) and the temporal random effect (rw1_month_all), with the latter two exhibiting minimal variability. In the top panel, the spatial random effect remains close to zero across all observed SPDE mesh vertices, with very narrow credibility intervals. This suggests that the variability introduced by this random effect is minimal, and that it does not contribute significantly to the outcome. The middle panel displays the IID random effect for individual neighbourhood, representing individual-level variability. Here, substantial fluctuations are observed, with several indices showing large deviations from zero, accompanied by wider credibility intervals for specific sections of the index. This finding highlights the significant role that the IID random effect associated with neighbourhood factors plays in the model, contributing considerable variability to the outcome. The steep changes in mean values for certain neighbourhoods highlight strong spatial heterogeneity, suggesting that specific neighborhoods possess unique characteristics that drive differences in the outcome. In the bottom panel, the random effect for rw1_month_all, which captures temporal variation across months, remains relatively stable around zero. However, the wider credibility intervals, particularly at the extremes of the plot, suggest uncertainty in the estimation of this effect. This suggests that while there is some variability over time, the overall effect is small, and the wide intervals indicate uncertainty in how the model captures monthly fluctuations. The wider band towards the ends may reflect more uncertainty in the estimation for early and later months, which could result from less data or greater variability in those periods.

In contrast, S5 Fig depicts the random effects for the spatiotemporal interaction model. This figure contains four panels, with the final plot representing the spatiotemporal interaction effect. Notably, the IID random effect remains consistent across both models. However, the credible intervals for the spatial and temporal random effects are wider in the spatiotemporal interaction model, though the variation remains close to zero. This indicates increased uncertainty in these random effects estimates but suggests that their overall contribution remains minimal. In the last panel, the spatiotemporal interaction effect displays minimal variation, with values remaining relatively stable around zero. This stability indicates that the interaction effect contributes little additional variability to the model, suggesting limited region-period-specific deviations beyond those captured by the main spatial and temporal effects.

For the spatiotemporal separable model, the spatial effect parameters of *κ* and τ have mean values of 53.70 and 0.29, respectively. These values represent the marginal posterior distributions of the two hyperparameters for the spatial random field. Using these parameters, we calculated the spatial range *r* to be 0.452 km, or 452 meters. In contrast, for the spatiotemporal interaction model, we found of *κ* to be 35.62 and of τ to be 1.149, resulting in a spatial range of 0.064 km, or 64 meters. Notably, the spatiotemporal separable model shows a larger spatial range and higher *κ* value compared to the interaction model, indicating a broader influence of spatial correlations. To further assess the goodness-of-fit of our best-fitting model, we evaluated its predictive performance at unsampled locations [53]. Fig 4 reports the comparison between predicted and observed COVID-19 infection. The pearson correlation coefficient is 97%, which indicates a strong positive association between the predicted and the observed COVID-19 infection.

**Fig 4.**
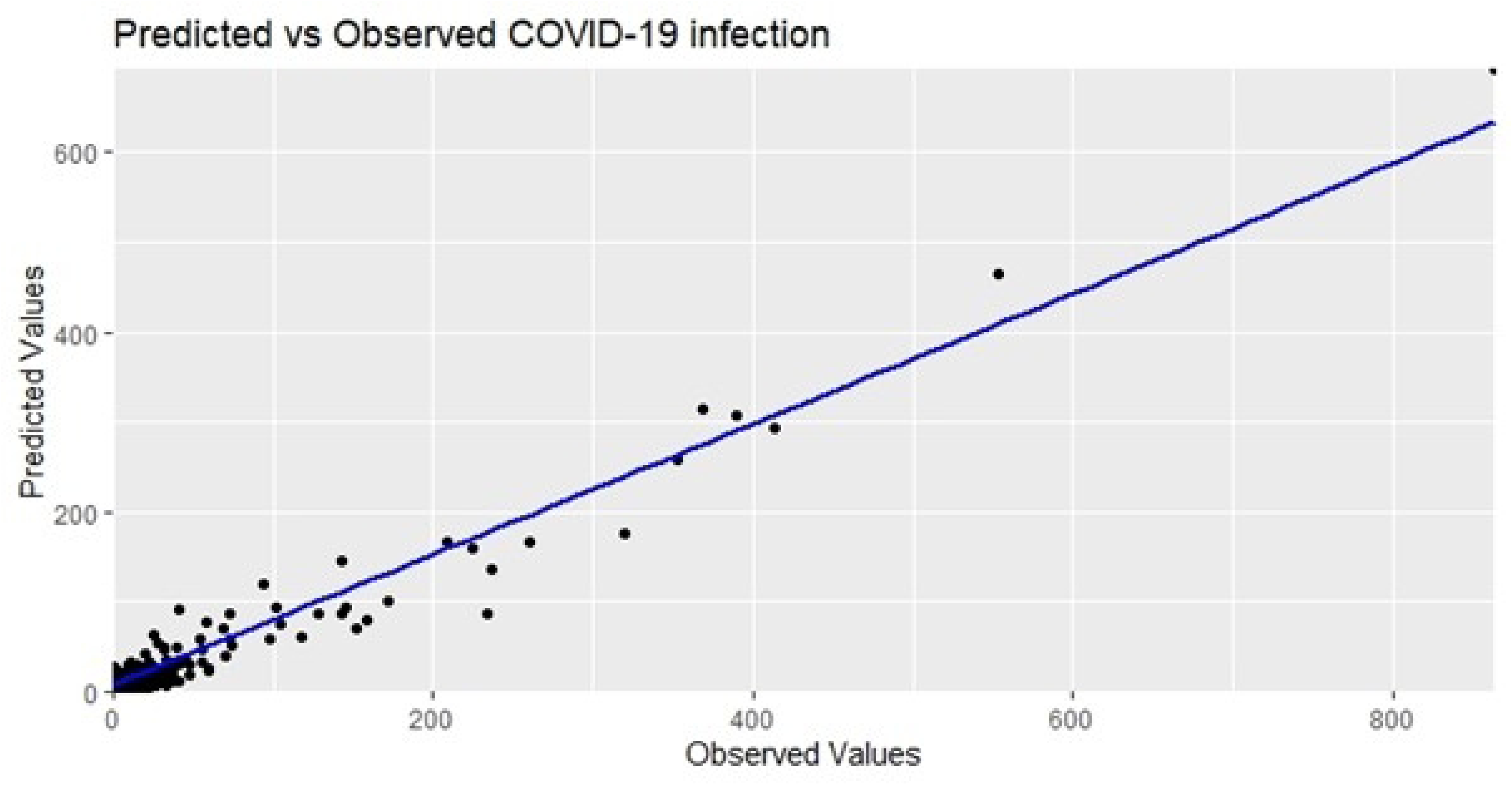
Comparison between the predicted and observed infection for the testing records.

A dynamic map that depicts the prediction from the spatiotemporal interaction model has been developed in an R-shiny app and accessible through this link (covid19_Dakar). A sample view of the shiny app is presented in S6 Fig. The risk of COVID-19 infection is higher in the western Dakar, particularly in the neighbourhoods of the following Communes: Fann-Point-E, Amitie, Plateau, Mermoz Sacre Coeur, Ngor, Sicap Liberte, Dieuppeul, Grand Yoff and Patte d’Oie. Central and Eastern peripheral areas have the lowest probabilities of COVID-19 infection.

## 4. Discussion

In this study, we developed Bayesian spatiotemporal models to examine the role of contextual risk factors in the spread of COVID-19 infection at the neighborhood level in Dakar. In addition to assessing the influence of these risk factors, we quantified the uncertainties associated with both the fixed effects of covariates and the random effects of spatial, temporal, and spatiotemporal components. A key outcome of this study is the mapping of areas at higher risk of COVID-19 infection in Dakar. Furthermore, the strong correlation between the predicted values from the model and the observed values underscores the high predictive accuracy of the best-fitted spatiotemporal interaction model.

To investigate the relationship between the covariates and COVID-19 infection, various contextual factors were considered. These factors were extracted from remote sensing images, which have proven useful for obtaining environmental and socioeconomic information. Remotely sensed socioeconomic and environmental variables have been widely applied to enhance our understanding of the local epidemiology of infectious diseases [54–59], including COVID-19 disease [60]. Population density is a very important feature of compact cities and an important proxy of socio-economic conditions along with land use characteristics such as built-up residential areas and built-up non-residential areas examined in this study. Regarding the contrasting statistical correlations that were previously found in the literature, showing a positive [10,14], then a negative correlation [5,8] between COVID-19 infection and population density, this study was an opportunity to further investigate this issue, by using very detailed data, and a more rigorous analytical approach. Our results confirm evidence of contrasting statistical correlations and highlight the impact of the scale of measurement on these correlations. On the one hand, COVID-19 infection was found to be significantly negatively associated with population density when no further adjustments were made and when population density was calculated using the total area of the spatial units as the denominator. On the other hand, when using adjusted population density, which considers only the built-up area as the denominator, a significant positive association with COVID-19 infection was observed. Therefore, adjusting population density before its application in our spatial statistical analysis has addressed issues related to the intrinsic properties of raw, non-adjusted population density such as the implicit assumption of uniformity over spatial units [13], the varying definitions of boundaries and the effect of varying scale of measurements [61]. Since the early stages of the pandemic, areas with high population density have experienced the greatest burden of COVID-19 infections [62,63]. Population density has significantly influenced the spatial distribution of COVID-19 [64], and has been a key indicator for implementing control measures such as physical distancing [65] that aimed to combat the spread of the disease. Our findings link negative correlation between COVID-19 infection and population density to the omission of important methodological considerations. Conversely, the positive association that emerges following further adjustements confirms the role of population density as a primary risk factor for infectious diseases [61], including COVID-19 diseases.

Similarly, we found a significant positive association between residential areas and COVID-19 infection. In contrast, COVID-19 infection was negatively associated with non-residential areas, which include ACS areas. The distance to marketplaces was found to be not significant, suggesting that the effect of mobility was not fully captured by the geospatial predictors used in this study. Despite the complexities surrounding the influence of neighborhood features on the spread of COVID-19 [66], the positive association between COVID-19 infection and residential areas provides insights into local exposure in relation to built-up areas and land use characteristics. Additional information about residential dwellers, such as household social composition and income levels, could further reveal significant disparities in the spread of COVID-19 across residential areas [67].

The spatiotemporal model with space-time interaction demonstrated higher predictive performance compared to the non-spatial model and the spatiotemporal model with separable space-time effects. Including the space-time interaction improved the model performance as it accounts for specific local spikes, which cannot be explained by overall spatial or temporal effects alone. Indeed, the spatiotemporal interaction term has the benefit of capturing localised deviations in the infection risk, which is particularly valuable for identifying transient, area-specific surges, thereby enhancing model’s sensitivity to local heterogeneity [40,41].

There is an important limitation that deserves to be highlighted. The epidemiological data provided by the Ministry of Health includes only the location of cases resulting from community transmission, therefore total case counts in some areas could be underrepresented. No information is available on the spatial distribution of other types of infection, such as the so-called “contact cases”. Including these in the analysis could have refined our results. Furthermore, despite the low mortality rate from COVID-19, data on COVID-19 related-deaths could have provided additionnal insigths into the role of contextual risk factors in the spread of the disease. In this study, we aggregated the number of confirmed infection by months, which in turn smooths the daily reporting noise. Nonetheless, missing data, under-reporting and uneven testing across neighbourhoods may still affect the results. For instance, areas with better testing access might report more cases, which could confound the association with contextual risk factors. Potential data biases may arise from incomplete reporting or spatial variations in the detection of the infection, although greater concentration of health facilities and shared testing infrastructure in Dakar may limit these differences. Lastly, it is worth mentioning that, although remotely-sensed variables are useful proxies that capture meaningful landscape characteristics, census or survey-based household-level crowding and socioeconomic variables are useful indicators that could further refine our spatiotemporal model, and improve our understanding on the influence of contextual risk factors on COVID-19 infection.

## 5. Conclusion

The COVID-19 related social, health and economic impact has been gradually lessened, thanks to various efforts notably the rapid deployment of various vaccines that prompted a high level of population immunity. But in the post pandemic era, the disease is still circulating and the end of the global health emergency has led to the emergence of a global health threat. The risk of COVID-19 infection, or re-infection persists, as new virus variants can emerge, causing new surges in cases and deaths.

From a public health perspective, the research findings gained from this study can be valuable resources for health systems in low- and middle-income countries, for geographically targeted interventions. These outputs can serve as support tools for the optimised distribution of the COVID-19 vaccine and the monitoring of the potential COVID-19 outbreaks. Testing or vaccination efforts should prioritize high-risk western neighbourhoods, or dense residential areas.

Furthermore, the model can be adapted for the study of other infectious diseases, including malaria, tuberculosis and other neglected tropical diseases. Here we can leverage the flexibility of Bayesian spatiotemporal models, which has gained more popularity due to their capacity to incorporate spatial and temporal dependencies, uncertainties and intricate interactions. Bayesian computing has long been a complex task, but in the era of big data and the rapid advancement of computer technology, the Bayesian framework has been enhanced enough to allow in-depth investigations of epidemiological data, and the study of different diseases with a different and more extensive set of regressors.

## Acknowledgment

This work was supported, in whole or in part, by the Gates Foundation under the GRID3 project. The conclusions and opinions expressed in this work are those of the author(s) alone and shall not be attributed to the Foundation. Under the grant conditions of the Foundation, a Creative Commons Attribution 4.0 License has already been assigned to the Author Accepted Manuscript version that might arise from this submission. Please note works submitted as a preprint have not undergone a peer review process.

## Data availability

The epidemiological data used in this study is publicly available through a request from the Ministry of Health. The geospatial predictors that we utilised to perform the analyses are freely available through the WorldpPop (https://www.worldpop.org/datacatalog/) and MAUPP data repositories (https://maupp.ulb.ac.be/page/grippa2018/).

## Code availability

All the code used to support this research, as well as the R-shiny app we developed to visualise the results are available in the following repository: https://github.com/assanenian/INLA_Bayesian_Spatiotemporal

## Supporting information

**S1 Fig. SPDE triangulation of the study area**

**S2 Fig. Marginal posterior distributions of covariate coefficients for the separable spatio-temporal model**

**S3 Fig. Marginal posterior distributions of covariate coefficients for the spatiotemporal interaction model**

**S4 Fig. Marginal posterior distribution of the random effects components for the separable spatiotemporal model. Top: Marginal posterior mean of the spatial random effect *S*(.)**

**Middle: Marginal posterior mean of the iid effect u_i_**

**Bottom: Marginal posterior mean of the temporal random effect γ_t_**

***S5 Fig. Marginal posterior distribution of the random effects components for the spatiotemporal interaction model. Top panel: Marginal posterior mean of the spatial random effect *S*(.)***

***Second panel: Marginal posterior mean of the iid effect u_i_***

***Third panel: Marginal posterior mean of the temporal random effect γ_t_***

***Bottom panel: Spatiotemporal interaction effect***

**S6 Fig. Overview of the R-Shiny App that visualizes the comparison between the observed and predicted COVID-19 prevalence.**

## Author contributions

Conceptualisation, methodology: ANG, MWT, AND; formal analysis and validation: AND and SC; data curation: MWT; Writing and editing: ANG, SC, NS, CL, SL, EC, CCN, AL, AJT, OY; supervision: AND.

